# Unravelling Somatic Symptoms in Pakistan’s Urban Slums: A Sex-Stratified Multilevel Exploration of Individual and Household Factors

**DOI:** 10.1101/2025.10.07.25337547

**Authors:** Maheen Sughra, Aisha Irum, Muhammad Ibrahim, Adnan Ahmad Khan

## Abstract

Mental health conditions in low- and middle-income countries (LMICs) often manifest as somatic symptoms—physical complaints lacking clear medical causes—that complicate diagnosis and care. This study examines the prevalence and determinants of somatic symptoms in Dhoke Hassu, a low-income urban settlement in Rawalpindi, Pakistan, with particular attention to gender and household dynamics. Using data from 782 adults (18–75 years), we applied sex-stratified multilevel logistic regression to explore individual and household-level predictors. Overall, 72% of participants reported somatic symptoms. Results revealed that household-level factors explained 18% of the variance among women but were not significant for men, underscoring the influence of family context on women’s psychosomatic health. Across both sexes, older age, hypertension, and medical service utilization were strongly associated with symptom reporting. Gender-specific patterns emerged: higher body mass index, access to technology, and a family history of mental illness were protective for men, while women’s symptoms were linked to household roles and relational positioning. These findings highlight the need for integrated, gender-sensitive approaches in primary care. Interventions should embed mental health screening in routine healthcare, leverage digital tools for men, and address relational and household stressors for women. By situating somatization within the Social Ecological Model, this study advances understanding of multi-level influences on mental health in Pakistan’s urban slums and provides actionable pathways for policy and practice.

## 1. Introduction

Mental health disorders are a major contributor to the global disease burden, yet their impact remains underestimated in low- and middle-income countries (LMICs). Nearly one in three adults in Pakistan experiences a mental health condition, yet mental distress often goes unrecognized and untreated, especially in low-resource urban slums where structural vulnerabilities heighten psychological burden(1). In such settings, mental health issues frequently present as somatic symptoms - physical complaints such as pain, fatigue, or shortness of breath that lack identifiable medical causes. This “somatization” affects around 26-35% of the general population worldwide, and poses significant challenges for diagnosis and treatment, worsen individual wellbeing and strain already overburdened health systems in LMIC, further limiting access to care(2, 3).

These global trends are reflected in South Asia where nearly one in four adult women experience a mental health condition, such as anxiety or depression, often expressed through somatic complaints (4). The prevalence may be higher as seen from a study from Khyber Pakhtunkhwa reporting somatic symptoms among 90% of women and 75% of men(4). The extant social pressures and stigma aggravate these conditions, particularly for women, limit their care seeking(5, 6). Research also shows that a complex confluence of factors such as age (6), socioeconomic status (7), and household mental illness risk (8) contributes to the likelihood of somatization, which is more common among women(7). The Social Ecological Model (SEM) provides a useful framework to understand how these interactions of individual, household, and environmental factors shape somatic symptom expression, particularly in marginalized contexts with dense populations, poor living conditions, and limited healthcare access (8).

Despite these trends, research on somatization in Pakistan remains limited, particularly studies that unpack household-level and gender-specific dynamics in informal urban settlements. This study seeks to address this gap in Dhoke Hassu, a poor urban settlement in Rawalpindi that has limited access to mental healthcare and signifies considerable ethnic diversity. Using sex-stratified multilevel logistic regression, we examine how factors such as age, socioeconomic stressors, health history, and household relationships contribute to somatic symptoms and somatization in the context of poor urban areas in LMIC such as Pakistan.

## 2. Methodology

### Study Design and Setting

The data used in this research were drawn from a cross-sectional study conducted in June 2023 in Dhoke Hassu, a densely populated urban slum in Rawalpindi, Pakistan, designed to assess the prevalence and distribution of noncommunicable disease (NCD) risk factors— including hypertension, diabetes, obesity, and mental health. The original data were collected through a complete household enumeration survey and anthropometric measurements by Research and Development Solutions (RADS) in collaboration with the Akhter Hameed Khan Foundation (AHKF). For the present analysis, we retrospectively accessed this dataset with approval and consent from the primary investigators of the original study. The final sample comprised 782 adults aged 18–75 years, who had provided informed consent at the time of data collection. Ethical approval and oversight were provided by the Institutional Review Board of RADS, in accordance with established standards for research involving human participants.

### Outcome Variable

The primary outcome was the presence of somatic symptoms, operationalized as a binary variable. Four items from the Patient Health Questionnaire (PHQ-9) were selected to capture somatic manifestations of psychological distress:

- Item 3: Trouble falling or staying asleep
- Item 4: Feeling tired or having little energy
- Item 5: Poor appetite or overeating
- Item 7: Trouble concentrating on things, such as reading the newspaper or watching television

Participants who endorsed any of these symptoms were classified as experiencing somatic symptoms. This approach is supported by Schier et al. (2012), who demonstrated that these specific PHQ-9 items effectively capture somatic expression of distress across diverse populations, with high internal consistency (Cronbach’s α = 0.82) (9).

### Independent Variables

This study examined both individual and household-level predictors of somatic symptoms. Individual-level variables included demographic factors (age, education, marital and employment status), health indicators (Body Mass Index, self-reported hypertension), psychosocial elements (violence exposure, self-rated life satisfaction), and technological access (availability of a smart phone). Household-level factors included family disease risk (categorized as low, moderate, or high based on reported familial health background), family mental health history, and Household role (participant’s relationship to the household head).

This hierarchical structure supported a multilevel analytical framework to assess both individual- and household-level determinants of somatic symptomatology, accounting for clustering at the household level.

### Data Analysis

This study employed a sex-stratified multilevel logistic regression framework to account for the hierarchical structure of the data, where individuals were nested within households. Stratification by sex allowed us to compare how individual-level and household-level factors differentially influence somatic symptom reporting among men and women. The multilevel approach accounted for both within-household and between-household variation, providing correct estimation of standard errors and enabling calculation of the proportion of variability attributable to clustering at the household level (10).

We began with descriptive statistics for all household and individual variables, followed by bivariate mixed-effects multilevel logistic regression analyses to examine crude associations between independent variable and somatic symptoms. Both fixed effects (individual-level variation) and random effects (household-level variation) were estimated. All predictors were standardized to facilitate interpretation across the sex-stratified analyses. The decision to conduct separate analyses for men and women was theoretically grounded in prior research demonstrating gender-differentiated patterns in somatic symptom expression in South Asian contexts.

To systematically assess the contribution of household versus individual predictors, we employed a stepwise modeling strategy with three sequential models:

- **Null Model (Empty Model):** A baseline model with no predictors, used to estimate the degree of clustering of somatic symptoms within households. Including this model ensures that subsequent effects are interpreted relative to baseline clustering.
- **Model I:** Incorporated only individual-level factors (e.g., age, health conditions, BMI, life satisfaction). This step isolates the influence of individual characteristics while controlling household clustering.
- **Model II:** Added household-level predictors (e.g., family illness risk, family mental health history, relationship to household head) to the individual-level factors. Comparing Model II with Model I allowed us to determine whether household characteristics explained additional variance in somatic symptom reporting beyond individual factors.

Model fit was evaluated using Akaike Information Criterion (AIC) and Bayesian Information Criterion (BIC), with lower values indicating improved fit. Likelihood-ratio tests were also applied to compare nested models and assess the statistical significance of household-level variance. This sequential approach provided a transparent way to isolate the respective contributions of individual and household factors across genders.

All analyses were conducted in STATA using mixed-effects logistic regression. Ethical approval for the study, including population selection and consent procedures, was obtained from the Institutional Review Board of RADS, which is accredited by the Office of Human Research Protections.

## 3. Results

### Descriptive Analysis

Of the 782 participants, 52% were women (Table 1). Of these, 72% reported experiencing somatic symptoms. The majority (54%) were between 18-34 years of age, while 25% were 45 year or above. Forty percent were heads of the households. Health related concerns were notable, 31% had hypertension, and 46% had utilized some medical care. Mental health risks were common in that 93% reported a family history of mental illness.

**Table 1:** Demographic and Health-related Characteristics of study Participants.

| **Characteristics** | **N** | **%** |
| --- | --- | --- |
| Experienced Somatic Symptoms in the past 2 weeks |  |  |
| Experienced No Somatic Symptoms | 318 | 27.8 |
| Experienced Somatic Symptoms | 565 | 72.2 |
| Age |  |  |
| Age Group 1(18-24) | 204 | 26.1 |
| Age Group 2(25-34) | 214 | 27.4 |
| Age Group 3(35-44) | 171 | 21.9 |
| Age Group 4(45-54) | 98 | 12.5 |
| Age Group 5(55-80) | 95 | 12.2 |
| Body Mass Index (BMI) |  |  |
| Underweight | 69 | 8.8 |
| Normal | 294 | 37.6 |
| Overweight | 210 | 26.9 |
| Obese | 209 | 26.7 |
| Education |  |  |
| No Education | 348 | 44.5 |
| Primary Education | 154 | 19.7 |
| Middle Education | 98 | 12.5 |
| Secondary Education | 91 | 11.6 |
| College Education | 72 | 9.21 |
| University | 5 | 0.6 |
| Other | 14 | 1.8 |
| Married Status |  |  |
| Married | 572 | 73.2 |
| Single | 167 | 21.4 |
| Other | 43 | 5.5 |
| Employment Status |  |  |
| Unemployed | 389 | 49.7 |
| Employed | 330 | 42.2 |
| Other | 64 | 8.2 |
| Life Satisfaction |  |  |
| Satisfied | 620 | 79.3 |
| Neutral | 96 | 12.3 |
| Dissatisfied | 66 | 8.4 |
| Diseases |  |  |
| Hypertension |  |  |
| Yes | 245 | 31.3 |
| No | 537 | 31.3 |
| Utilization of Medical Services |  |  |
| Yes | 355 | 45.5 |
| No | 426 | 54.6 |
| Access to Technology |  |  |
| Has a Smartphone | 230 | 29.4 |
| Has a Phone | 230 | 29.4 |
| Does not have | 322 | 41.2 |
| Has been exposed to Violence |  |  |
| Never | 755 | 97.9 |
| Rarely | 11 | 1.4 |
| Sometimes | 4 | 0.5 |
| Often | 1 | 0.1 |
| Family Mental Health History |  |  |
| No Risk of Mental Illness in Family | 58 | 7.4 |
| Risk of Mental Illness in Family | 724 | 92.6 |
| Family Illness History Risk |  |  |
| Minimal Family History | 85 | 10.9 |
| Moderate Family History | 310 | 39.6 |
| High Family History | 387 | 49.5 |
| Relationship with Household Head |  |  |
| Head | 241 | 40.2 |
| Immediate Family | 182 | 30.3 |
| Extended Family | 166 | 27.7 |
| Other Relatives | 11 | 1.8 |

**Table 2:** Multilevel Logistic Regression Results for Psychosomatic Symptoms.

| Predictor | Women |  |  | Men |  |  |
| --- | --- | --- | --- | --- | --- | --- |
|  | Model 1:<br>Empty<br>Model | Model 2:<br>Individual<br>Variables<br>Only | Model 3:<br>Model 2 +<br>Household<br>Level<br>Variables | Model 1:<br>Empty<br>Model | Model 2:<br>Individual<br>Variables<br>Only | Model 3:<br>Model 2 +<br>Household<br>Level<br>Variables |
| Fixed Effects |  |  |  |  |  |  |
| Age |  |  |  |  |  |  |
| Age Group 2 |  | 1.77 | 1.48 |  | 1.77 | 2.05** |
| Age Group 3 |  | 1.84 | 1.51 |  | 2.70* | 3.64* |
| Age Group 4 |  | 3.16 | 2.44 |  | 3.85* | 5.15* |
| Age Group 5 |  | 2.31 | 2.30 |  | 1.80 | 2.64** |
| BMI |  | 0.88 | 0.94 |  | 0.75* | 0.74* |
| Education |  | 0.98 | 0.99 |  | 1.02 | 1.04 |
| Married<br>Status |  | 0.74 | 0.58 |  | 1.40 | 1.31 |
| Employment<br>Status |  | 1.13 | 1.11 |  | 1.23 | 1.25 |
| Life<br>satisfaction |  | 1.46 | 1.43 |  | 0.79 | 0.81 |
| Diseases |  |  |  |  |  |  |
| Hypertension |  | 2.70* | 2.55* |  | 2.71* | 2.64* |
| Utilization of<br>Services |  | 2.27* | 2.35* |  | 2.36* | 2.41* |
| Access to<br>Technology |  | 1.20 | 1.18 |  | 0.71** | 0.69** |
| Exposure to<br>Violence |  | 0.32 | 0.35 |  | 0.93 | 0.88 |
| Family Mental Health History |  |  | 0.45 |  |  | 0.25* |
| Family History Risk |  |  |  |  |  |  |
| Moderate Risk |  |  | 1.34 |  |  | 0.60 |
| High Risk |  |  | 0.78 |  |  | 0.54 |
| Relationship with Household Head |  |  | 0.57** |  |  | 1.35 |
| Random Effects |  |  |  |  |  |  |
| Household ID Variance | 0.95* | 0.61** | 0.71** | 0.12 | 0.21 | 0.25 |
| Intraclass Correlation (ICC) |  |  |  |  |  |  |
| ICC | .22=22% | .16= 16% | .18= 18% | .14=14% | 0.59= 5.9% | 0.72= 7.2% |
| Model Fit Statistics |  |  |  |  |  |  |
| AIC | 339.26 | 329.03 | 329.11 | 515.78 | 495.06 | 495.24 |
| BIC | 347.26 | 388.76 | 400.7765 | 523.66 | 553.97 | 565.93 |

Multilevel logistic regression results showed clear gender differences. Among women, household-level factors alone explained 22% of the variance in somatic symptoms. This decreased to 16% when individual-level variables such as age, health conditions, and lifestyle were added, but remained notable at 18% even after incorporating additional household characteristics. This indicates that household context plays an important and persistent role in shaping women’s psychosomatic health. In contrast, for men, household-level variance was not statistically significant, suggesting that somatic symptoms in men are primarily shaped by individual-level characteristics rather than family environment.

Across both sexes, older age, hypertension, and medical service utilization significantly increased the likelihood of reporting somatic symptoms. Among women, having a weaker relationship with the household head was associated with lower odds of symptoms. For men, several protective associations emerged: a higher body mass index (OR = 0.74) was linked to *reduced* odds of somatic symptoms, as was access to technology (OR = 0.69) and having a family history of mental health issues (OR = 0.25). These odds ratios below 1 indicate protective rather than risk-enhancing effects. Education, marital status, employment, and life satisfaction did not show significant associations with symptoms for either gender. Model fit statistics confirmed that including individual predictors substantially improved explanatory power, particularly for men, reinforcing that men’s somatic symptoms are better explained by personal health and resource factors, while women’s symptoms remain strongly shaped by the household context.

## 4. Discussion

Gender differences were central to our findings: household-level variation was substantially greater among women (ICC = 18%) than men, indicating that women’s somatic symptoms are more strongly shaped by family context and relational dynamics, while men’s symptoms are primarily determined by individual factors. Overall prevalence was high (72%), with older age, hypertension, and higher utilization of medical services consistently linked to greater symptom reporting in both sexes. Gender-specific associations further highlighted this divide: among men, higher BMI (OR = 0.75), access to technology (OR = 0.71), and a family history of mental illness (OR = 0.25) were protective, whereas among women, a weaker role as household head (OR = 0.57) was linked to lower symptom reporting. These patterns reinforce that men’s psychosomatic health is largely influenced by personal health and resources, while women’s is more deeply embedded in household structures and gendered roles.

Household-level variation was significantly prominent among females (0.71, ICC=18%), suggesting that household dynamics play a greater role in shaping psychosomatic health for women compared to men. This finding aligns with the Social Ecological Model (SEM), which emphasizes the importance of interpersonal and household-level influences on individual health outcomes(8). In patriarchal and resource-constrained settings, women often face greater psychosocial burdens related to caregiving roles, limited autonomy, and relational stress—all of which may intensify somatic symptom expression. SEM provides a useful lens for understanding how these multi-layered influences manifest differently across genders, reinforcing the need to look beyond individual-level risk factors when addressing women’s mental health in slum communities. Supporting this perspective, a systematic review by Abdi et al. (2021) highlighted that among women living in slum areas, gender and household-level factors were significantly associated with poorer mental health outcomes, emphasizing the need for multi-level interventions that address both individual and structural determinants(11).

Multiple studies indicate that women often exhibit and report more somatic symptoms than men across cultures(12). Women tend to present with higher counts and intensity of bodily complaints, sometimes reflecting underlying distress that might be underreported by men. In low- and middle-income countries (LMICs), this pattern persists. (13). The propensity for women to experience and report such symptoms more frequently may be tied to gender roles and norms. Women’s greater willingness or need to report physical symptoms – whether due to socialization or greater exposure to certain stressors – contributes to higher diagnosed rates of somatic symptom disorders in women(14). This may reflect the psychological burden of caretaking roles, family expectations, and power imbalances that women often face in patriarchal household structures. Men, on the other hand, might under-report or differently contextualize somatic complaints, potentially leading to under-recognition of their psychosomatic distress. In contrast, men’s somatic symptoms were more closely linked to personal factors such as BMI and access to technology, suggesting that men may experience or report distress through health-related autonomy or constraints, rather than family dynamics.

Subsequently, the consistent association between hypertension, older age, and somatic symptoms across both sexes highlights the deep interconnection between chronic physical conditions and psychological distress. Epidemiological data from primary care in LMICs show that patients with chronic medical conditions have a significantly greater co-occurrence of common mental disorders than physically healthy patients (15). For instance, one survey noted that the prevalence of diagnosable psychological disorders was about 57% in patients with chronic somatic diseases, compared to 49% in those without chronic conditions. Utilization of healthcare services by individuals reporting somatic symptoms may reflect ongoing search for physical explanations to what may be, at least in part, psychological distress. This pattern is common in low-resource settings were mental health stigma and lack of awareness limit emotional expression, especially among women (16).

Notably, access to technology was a protective factor for men. Men often face strong stigma against expressing vulnerability or seeking help for mental health issues through traditional means. Online platforms and smartphone apps provide a discreet avenue for men to learn about mental health, assess their own symptoms, and even engage in counseling or peer support without the fear of public exposure. This is also aligned with the findings from a study which highlighted that mental health service delivery in rural Pakistan was directly influence by internet connectivity (17). Access to digital platforms may serve as a crucial mental health resource, especially for men who may prefer seeking help or information privately.

## 5. Conclusion

This study revealed a high prevalence of somatic symptoms among adults in an urban slum in Pakistan and demonstrated that both individual and household-level factors contribute to this burden in gender-specific ways. While men’s symptoms were more closely linked to physical health and access to personal resources, women’s symptoms appeared deeply influenced by family dynamics and social roles.

## Data Availability

The data supporting this study are held by Research and Development Solutions (RADS) and are available upon reasonable request. Interested researchers may contact RADS at to request access.

## 6. Policy Implications

Given the high burden of somatic symptoms and their connection to both chronic physical and psychological health, there is a pressing need to integrate early mental health screening into primary care. Primary care providers often serve as the first point of contact for individuals presenting unexplained physical symptoms. Embedding routine mental health assessments-especially for patients with chronic conditions like hypertension-can help uncover underlying emotional distress and reduce unnecessary medical consultations.

Policy efforts in Pakistan should consider gender-sensitive approaches. For women, community-level outreach initiatives that address family-based stressors and gendered expectations can help reduce psychosomatic distress. For men, technology-based interventions such as mobile mental health applications or workplace wellness programs can serve as accessible platforms for support. Collaborative care models involving both physical and mental health professionals have shown success in similar low-and middle-income contexts (Whitfield et al., 2023) and should be adapted for slum populations in Pakistan.

## 7. Limitations

This study has several limitations. First, it uses cross-sectional data, which limits causal interpretation. Second, the measurement of somatic symptoms relied on self-reported data, which may be subject to reporting biases. Third, while the sample was drawn from an urban slum, the findings may not be generalizable to rural settings or other urban populations in Pakistan.

## 8. Directions for Future Research

Future research should consider longitudinal designs to better understand causal pathways between psychosocial factors and somatic symptoms. Additionally, qualitative studies could explore how gendered roles and household dynamics are perceived by individuals with somatic symptoms, enriching the interpretation of quantitative findings.

---

**Figure 1:**
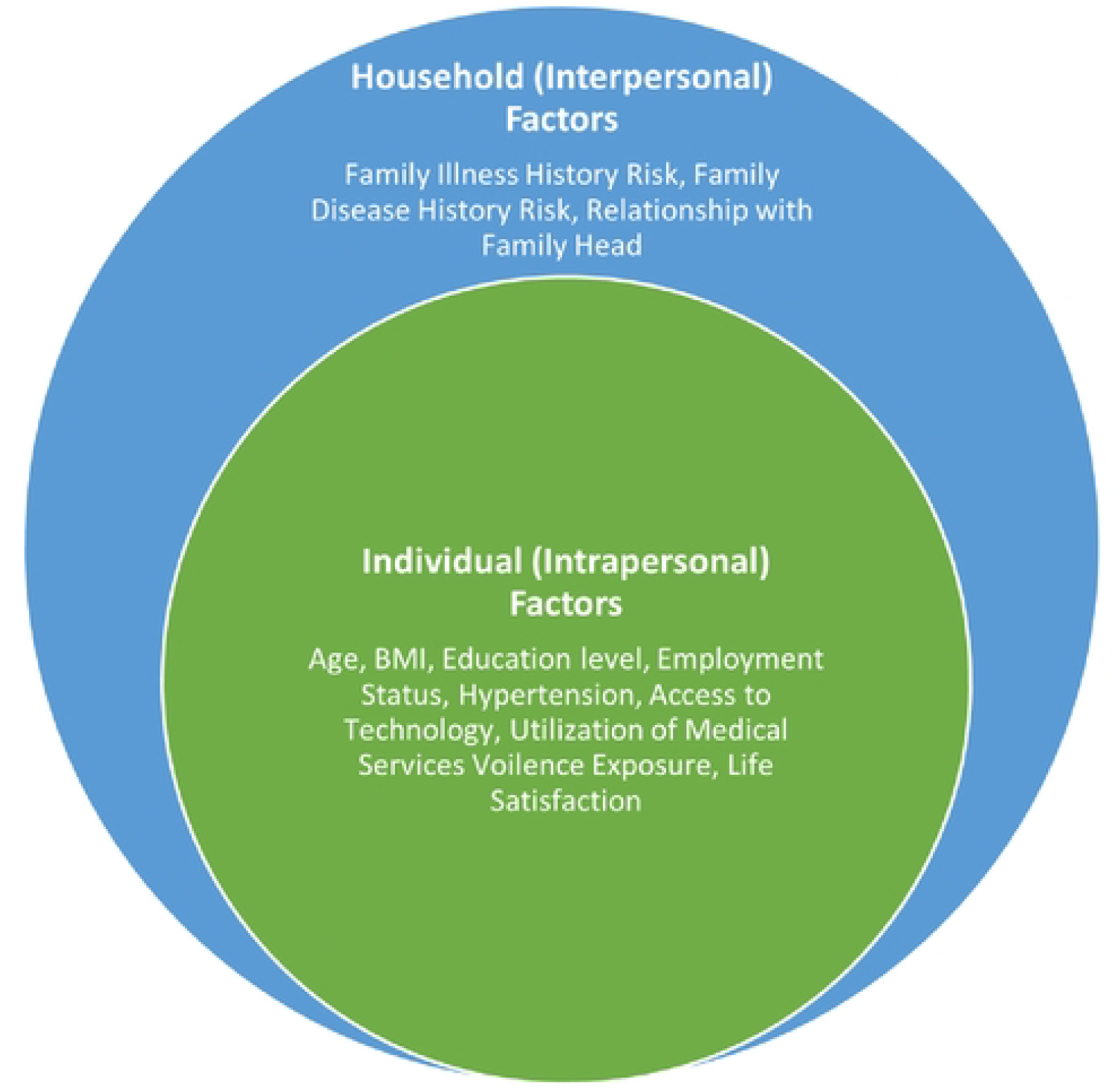
Multilevel Relationship of Somatic Symptoms in Urban Slum of Pakistan based on the Social Ecological Model {SEM}.

